# Genetic risk of inflammatory bowel disease is associated with disease course severity

**DOI:** 10.1101/2024.11.01.24316569

**Authors:** Marie Vibeke Vestergaard, Kristine Højgaard Allin, Anne Krogh Nøhr, Jesus Vicente Torresano Lominchar, Heidi Søgaard Christensen, Henrik Krarup, Kaspar René Nielsen, Henrik Albæk Jacobsen, Jonas Bybjerg-Grauholm, Marie Bækvad-Hansen, Rasmus Froberg Brøndum, Martin Bøgsted, Lone Larsen, Aleksejs Sazonovs, Tine Jess

## Abstract

**Background and aims:** Genetic susceptibility to inflammatory bowel disease (IBD) has been widely studied, whereas the genetic contribution to disease progression over time remains relatively under-investigated. In a Danish nationwide cohort, we aimed to explore if genetic susceptibility to IBD is associated with disease severity.

**Methods:** We estimated polygenic scores (PGS) for IBD susceptibility for 3,732 patients with Crohn’s disease (CD), 4,535 patients with ulcerative colitis (UC), and 9,469 controls, and investigated their association with disease outcomes, including inflammatory markers, hospitalizations, major surgeries, and medication use. A composite severity outcome was defined based on the first three years following diagnosis. Lastly, we evaluated disease extent as a possible mediator of the association.

**Results:** Increased susceptibility PGS was associated with higher fecal calprotectin and C-reactive protein levels, and decreased hemoglobin levels. When comparing the highest versus lowest PGS quintile, we observed a hazard ratio (HR) for major surgery of 2.74 (*P*=7.19×10^-18^) in patients with CD and of 2.04 (*P*=4.36×10^-7^) in UC. Patients with severe disease had higher susceptibility PGS than patients with less severe disease (CD: OR=1.25, *P*=3.36×10^-9^; UC: OR=1.33, *P*=1.40×10^-15^ per SD increase in PGS). Additionally, PGS was associated with a higher need for corticosteroids, immunomodulators, and biologic therapies. Adjusting for disease extent reduced the estimated associations for CD but had little impact on observed associations for UC.

**Conclusion:** Patients with higher genetic burden for developing IBD also experience a more severe disease course. For patients with CD this link was largely mediated by disease extent, however, this was not the case for UC, which suggests a shared genetic architecture between disease susceptibility and severity.

## Introduction

Inflammatory bowel disease (IBD) is a chronic, progressive, immune-mediated intestinal disease with an increasing incidence and prevalence globally. The two most common subtypes of the disease are Crohn’s disease (CD) and ulcerative colitis (UC).^1,2^ Patients are often diagnosed in early adulthood and experience a serious, life-altering condition that does not currently have curative treatment. The disease course is highly heterogeneous, ranging from relatively mild symptoms to severe disease characterized by uncontrolled gastrointestinal inflammation and repeated hospitalizations and surgeries.^3,4^ A better understanding of which patients will experience a more severe disease course would be the first step to improve treatment of this subgroup. However, our current limited understanding of IBD and the factors driving its heterogenicity make it difficult to anticipate a patient’s disease course.

Genome-wide association studies (GWAS) of IBD have primarily focused on disease onset by comparing patients with IBD to population controls. To date, more than 400 susceptibility loci have been identified.^5,6^ However, research on the genetic contribution to disease progression is more limited. In an analysis of approximately 2700 patients with CD performed by Lee *et al*.,^7^ four individual loci were associated with the need for immunomodulators or gastrointestinal surgeries. These loci were not associated with disease susceptibility.

Polygenic scores (PGS) use the results from GWAS to calculate the aggregated effect of trait-associated variants carried by an individual. To date, PGS has primarily been used to model the individual risk of disease onset for complex diseases where a well-powered, large-scale GWAS exists. For IBD, PGS can predict disease onset with limited performance (area under the receiver operating characteristic curve (AUC) = 0.63) and only identifies ∼3.2% of the population with a threefold increased risk of IBD.^8^ As for genetics overall, very little is known about the impact of genetics on the course of IBD. Although a strong association between PGS and disease extent has been reported,^9^ previous studies have failed to identify an association between susceptibility PGS and anti-TNF response,^10^ and CD prognosis stratified by disease extent.^7^

In this study, we used a large, unique population-based IBD cohort with both genetics and long-term clinical data to investigate the hypothesis that an individual’s genetic burden, as measured by their IBD susceptibility PGS, impacts the severity of the disease course.

## Methods and materials

### Danish registries

The study was based on data from Danish registries, including the Danish Civil Registration System,^11^ The Danish National Patient Registry (DNPR),^12^ The Danish National Prescription Registry,^13^ The Danish nationwide Registry of Laboratory Results for Research (RLRR)^14^ and the Danish Pathology Registry (DPR)^15^. Combined, they hold information on inpatient hospital contacts, International Classification of Diseases 8^th^ and 10^th^ revision (ICD-8 and -10) codes, surgical and other procedures since 1977, outpatient contacts since 1995, redeemed prescriptions for Danish residents at all pharmacies since 1994, results of biochemistry and hematological tests from hospitals and general practitioners since 2008 with complete coverage from 2015, and diagnostic statements from pathology departments since 1997. We had data available until September 2022. All registry codes used are listed in Table S1.

### Source populations

#### The PREDICT neonatal blood spot cohort (PREDICT-NBS cohort)

The Danish National Biobank stores neonatal blood spots from almost all Danes born since 1982.^16^ The PREDICT-NBS cohort includes neonatal blood spots from individuals born and diagnosed with IBD between April 1981 and December 2018.

Identification of patients with IBD was based on previously described criteria.^2^ IBD patients should have at least two IBD-related hospital registrations within a two-year period, or two separate inpatient contacts recorded in the DNPR. Assignment of CD or UC was based on the two most recent hospital contacts if these were concordant, and otherwise according to the highest number of diagnoses. Date of diagnosis was the date of the first IBD-related hospital registration. Patients not living in Denmark during the two years before this first IBD-related hospital registration were excluded to ensure a valid date of diagnosis. Each IBD patient was matched 1:1 to controls based on sex, date of birth, and no diagnosis of IBD prior to the patient’s diagnosis date. Matched controls that developed IBD from end of cohort selection (December 2018) to end of available registry data (September 2022) were considered IBD patients.

#### The North Denmark IBD cohort (NorDIBD)

The NorDIBD cohort is a population-based cohort of all patients from the North Denmark Region with a confirmed IBD diagnosis from 1978 to 2020.^17^ From this cohort, 940 patients with IBD and a further 973 blood donors with no IBD diagnosis were genotyped from whole blood. The date of IBD diagnosis was based on patient health records.

Overlapping individuals between the two cohorts (n=233) were randomly excluded from one of the cohorts, and only individuals of European ancestry were included. Details on genotyping, quality control, and generation of genetic principal components (PCs) were previously described^18^ and available in Supplement 1 and summarized in Figure S1.

### Polygenic score

We downloaded and applied variant loadings for calculating PGS for CD and UC susceptibility generated by Middha *et al.*^19^, to our dataset. These variant loadings were derived from summary statistics from the IIBDGC^20^ and calibrated on UK Biobank data. We calculated PGS using Plink; 738,417 out of 744,682 (>99%) and 738,314 out of 744,575 (>99%) available variants were included for calculating CD and UC susceptibility PGS, respectively. We also calculated CD and UC susceptibility PGS for the controls. The mean (SD) of CD PGS across controls and CD patients was 1.00 (1.27) and the corresponding mean (SD) of UC PGS across controls and UC patients was -0.95 (1.01). The PGS scores were normalized to standard deviations (SDs) from the mean to improve interpretability.

### Outcomes

We examined the association between susceptibility PGS and markers of inflammation i.e., C-reactive protein (CRP), fecal calprotectin (f-calpro) and hemoglobin at time of diagnosis and during follow-up. At diagnosis was defined as ± six months from the date of diagnosis, and the post-diagnosis phase was divided into five one-year intervals from half a year after diagnosis ((0.5,1.5), [1.5, 2.5), [2.5,3.5), [3.5,4.5), and [4.5,5.5) years).

Our primary outcomes were time to IBD-related hospitalization exceeding two days and time to major IBD-related surgery. We excluded patients diagnosed before January 1, 1996, to ensure completeness of relevant registries. This resulted in a final study population of 3732 patients with CD (3354 from the PREDICT-NBS cohort and 378 from the NorDIBD cohort), and 4535 patients with UC (4173 from the PREDICT-NBS cohort and 362 from the NorDIBD cohort) (Figure S2).

For our secondary outcome we excluded patients with less than three years of follow-up (CD=44, UC=44) to enable the classification of patients as having a severe versus less severe disease course based on the first three years following diagnosis. For patients with CD, we defined a severe disease course as experiencing I) two or more IBD-related hospitalizations exceeding two days, II) two or more IBD-related major surgeries, III) one IBD-related hospitalization and one IBD-related major surgery not overlapping in time, or IV) a total use of at least 5000 mg prednisolone equivalent systemic corticosteroids within the first three years following diagnosis (Figure S3). For patients with UC the severity definition was modified to experiencing I) two or more IBD-related hospitalizations exceeding two days, II) one or more IBD-related major surgeries, or III) a total use of at least 5000 mg prednisolone equivalent systemic corticosteroids. Patients not fulfilling these criteria were considered less severe.

We further tested each entity of the composite severity outcome separately: two or more IBD-related hospitalizations exceeding two days, two or more IBD-related major surgeries, one or more IBD-related major surgeries, and a total use of at least 5000 mg systemic corticosteroids within the first three years following diagnosis. Additionally, we evaluated the association with treatment with biologics, and immunomodulators for patients diagnosed after January 1, 2003, and the cumulative use of different treatment options within the first three years.

Definitions and codes used are listed in Table S1.

## Statistical analyses

At- and post-diagnosis biochemistry test values of CRP, f-calpro, and hemoglobin were evaluated for association with susceptibility PGS using a linear regression model adjusting for the covariates sex, cohort, calendar year, age at diagnosis, and the first ten PCs. This was only done for individuals with available measurements. Where there were multiple samples per individual in the same interval, the sample closest to date of diagnosis was chosen for the at-diagnosis interval, whereas the median value was chosen for the post-diagnosis intervals. Test values were transformed using log transformation for CRP and f-calpro to approach a Gaussian distribution.

We analyzed the association between susceptibility PGS and time to first IBD-related hospitalization exceeding two days and first major IBD-related surgery using Cox proportional hazards regression models. Censoring was based on death, emigration, or end of study. The model included adjustments for the above-mentioned covariates. Cumulative incidence curves were generated to visualize the association by dividing patients into PGS quintiles.

We analyzed the association between susceptibility PGS and our primary and secondary binary outcomes using logistic regression, adjusting for the above-mentioned covariates.

Reported p-values from tests of secondary outcomes and from biochemistry test values were adjusted for multiple testing using the Bonferroni correction. All results are reported per SD increase of susceptibility PGS.

## Sensitivity analyses

Sensitivity analyses were generated to evaluate the disease extent as a mediator of the association between susceptibility PGS and disease course severity.

We repeated the statistical analyses after recalculating the PGS by excluding the genetic loci MST1, NOD2, ATG16L1, and the HLA region ±500 kb, as these genetic regions are known to associate with disease extent.^9,21^

As a final sensitivity analysis, we inferred disease extent by using information from the DPR for the subset of patients with available data^22^. Ileal CD (L1) was inferred if patients had pathology registrations in ileum but not colon, colonic CD (L2) if there were registrations in the colon but not ileum, and ileocolonic CD (L3) for registrations in both the ileum and colon. Upper disease CD (L4) was coded as a separate parameter based on registrations in the upper gastrointestinal tract. Ulcerative proctitis (E1) was inferred if patients had registrations in the rectum but not colon, left-sided colitis (E2) if there were registrations in the descending colon to the sigmoid colon, and extensive colitis (E3) if there were registrations in the cecum to the transverse colon. Data was extracted from 30 days before to three years after diagnosis. If there were multiple records, the most widespread disease extent was selected. Selecting patients with inferred disease extent limited the cohort to 1970 patients with CD and 3067 patients with UC. Disease extent was included as a covariate when repeating the analyses.

## Ethical approval

The PREDICT-NBS cohort was approved by the Research Ethical Committee of the Capital Region of Denmark (H-20048987) and no direct consent was required. Collecting and analyzing blood samples from the NorDIBD cohort were approved by the Research Ethical Committee of the North Denmark Region (N-20170005-N-20200071). Oral and written consents were obtained, and we followed up on whether any participant withdrew their consent.

## Results

We included 8,267 patients with IBD (3,732 (45%) patients with CD and 4,535 (55%) with UC) and 9,469 control individuals (Table 1). Among patients with CD, the mean age at diagnosis was 21.8 years (SD=7.7), and 56% were female. Among patients with UC, the mean age at diagnosis was 23.3 years (SD=8.4), and 53% were female. Control individuals had a mean normalized PGS of -0.221 for CD PGS and -0.235 for UC PGS. Patients with CD had a mean normalized CD PGS of 0.528, whereas patients with UC had a mean normalized UC PGS of 0.469.

**Table 1:** Baseline characteristics of study population. Numbers (percentage) are reported in the table.

|  | All<br>N = 8267 (100) |  | The PREDICT-NBS cohort<br>N = 7527 (91.0) |  | The NorDIBD cohort<br>N = 740 (9.0) |  |
| --- | --- | --- | --- | --- | --- | --- |
|  | CD | UC | CD | UC | CD | UC |
| **N** | 3732 (45.1) | 4535 (54.9) | 3354 (44.6) | 4173 (55.4) | 378 (51.1) | 362 (48.9) |
| **Sex** |  |  |  |  |  |  |
| Female | 2081 (55.8) | 2420 (53.4) | 1882 (56.1) | 2221 (53.2) | 199 (52.6) | 199 (55.0) |
| Male | 1651 (44.2) | 2115 (46.6) | 1472 (43.9) | 1952 (46.8) | 179 (47.4) | 163 (45.0) |
| **Age at diagnosis** |  |  |  |  |  |  |
| <17 years | 823 (22.1) | 774 (17.1) | 784 (23.4) | 756 (18.1) | 39 (10.3) | 18 (5.0) |
| 17-40 years | 2822 (75.6) | 3615 (79.7) | 2570 (76.6) | 3417 (81.9) | 252 (66.7) | 198 (54.7) |
| ≥40 years | 87 (2.3) | 146 (3.2) | - | - | 87 (23.0) | 146 (40.3) |
| **Calendar year at diagnosis** |  |  |  |  |  |  |
| 1996-2005 | 534 (14.3) | 579 (12.8) | 434 (12.9) | 491 (11.8) | 101 (26.7) | 88 (24.3) |
| 2005-2015 | 1943 (52.1) | 2474 (54.6) | 1790 (53.4) | 2312 (55.4) | 153 (40.5) | 162 (44.8) |
| 2015-2020 | 1254 (33.6) | 1482 (32.7) | 1130 (33.7) | 1370 (32.8) | 124 (32.8) | 112 (30.9) |

CD: Crohn's disease, UC: ulcerative colitis, PREDICT-NBS: PREDICT neonatal blood spots, NorDIBD: North Denmark inflammatory bowel disease

### Biochemistry profile is affected by susceptibility genetics

Increased susceptibility PGS was associated with disease activity markers of gastrointestinal and systemic inflammation (Figure 1). At diagnosis, f-calpro was increased (CD: β=0.27, 95% CI=0.21-0.34 log(mg/kg) per SD increase, UC: β=0.21, 95% CI=0.13-0.28 log(mg/kg) per SD increase) and hemoglobin decreased (CD: β=-0.11, 95% CI=-0.15--0.07 mmol/L per SD increase, UC: β=-0.08, 95% CI=-0.13--0.04 mmol/L per SD increase) with higher PGS, whereas CRP was only increased with higher PGS for patients with CD (β=0.15, 95% CI=0.09-0.21 log(mg/L) per SD increase). For patients with CD, increased f-calpro levels remained significantly associated with higher PGS during the following disease course. Thus, we decided to further investigate the association between susceptibility genetics and disease course outcomes of patients.

**Figure 1:**
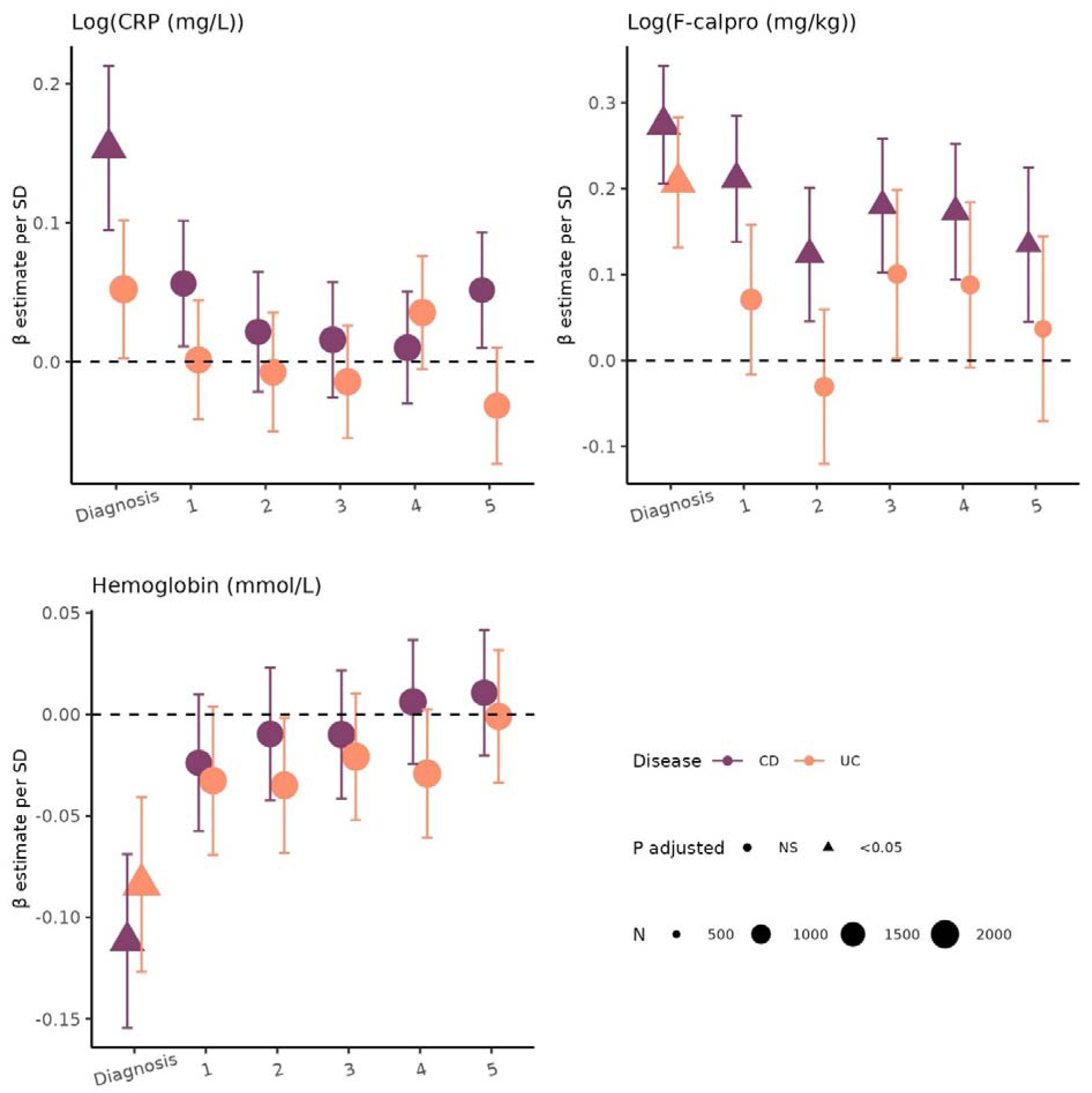
Association between susceptibility PGS and laboratory biochemical test results at different time points. Plot showing β estimates and their 95% CI for association between laboratory test results (CRP, f-calpro, and hemoglobin) and SD standardized susceptibility PGS for CD and UC separately. For each individual, the test results were calculated as following: Diagnosis = test result closest to date of diagnosis (±six months), and post-diagnosis = the median value of all tests in the time intervals (0.5,1.5), [1.5, 2.5), [2.5,3.5), [3.5,4.5), and [4.5,5.5) years after diagnosis. Estimates and 95% CIs are showed. *P*-values were adjusted for multiple testing using the Bonferroni correction for six tests. The dots are shaped based on significance after adjusting for multiple testing. Dot sizes are based on number of individuals with available test results in that given period. CD: Crohn’s disease, UC: ulcerative colitis, PGS: polygenic score, SD: standard deviation, CRP: C-reactive protein, f-calpro: fecal calprotectin, CI: confidence interval, NS: not significant

### Higher genetic burden of IBD susceptibility is associated with severe outcomes

We separated the CD and UC cohorts into quintiles based on susceptibility PGS. Looking into the time to first IBD-related hospitalization exceeding two days and time to major IBD-related surgery revealed cumulative incidence curves ranking based on their PGS quintile, where the quintile with lowest PGS had the longest event-free time (Figure 2). This pattern was consistent for both time to hospitalization and time to major surgery for both CD and UC. Comparing the highest PGS quintile to the lowest revealed a quicker time to hospitalization (CD: hazard ratio [HR]=1.81, 95% confidence interval [CI]=1.57-2.09, UC: HR=1.69, 95% CI=1.47-1.95) and to major surgery (CD: HR=2.74, 95% CI=2.18-3.45, UC: HR=2.04, 95% CI=1.55-2.69).

**Figure 2:**
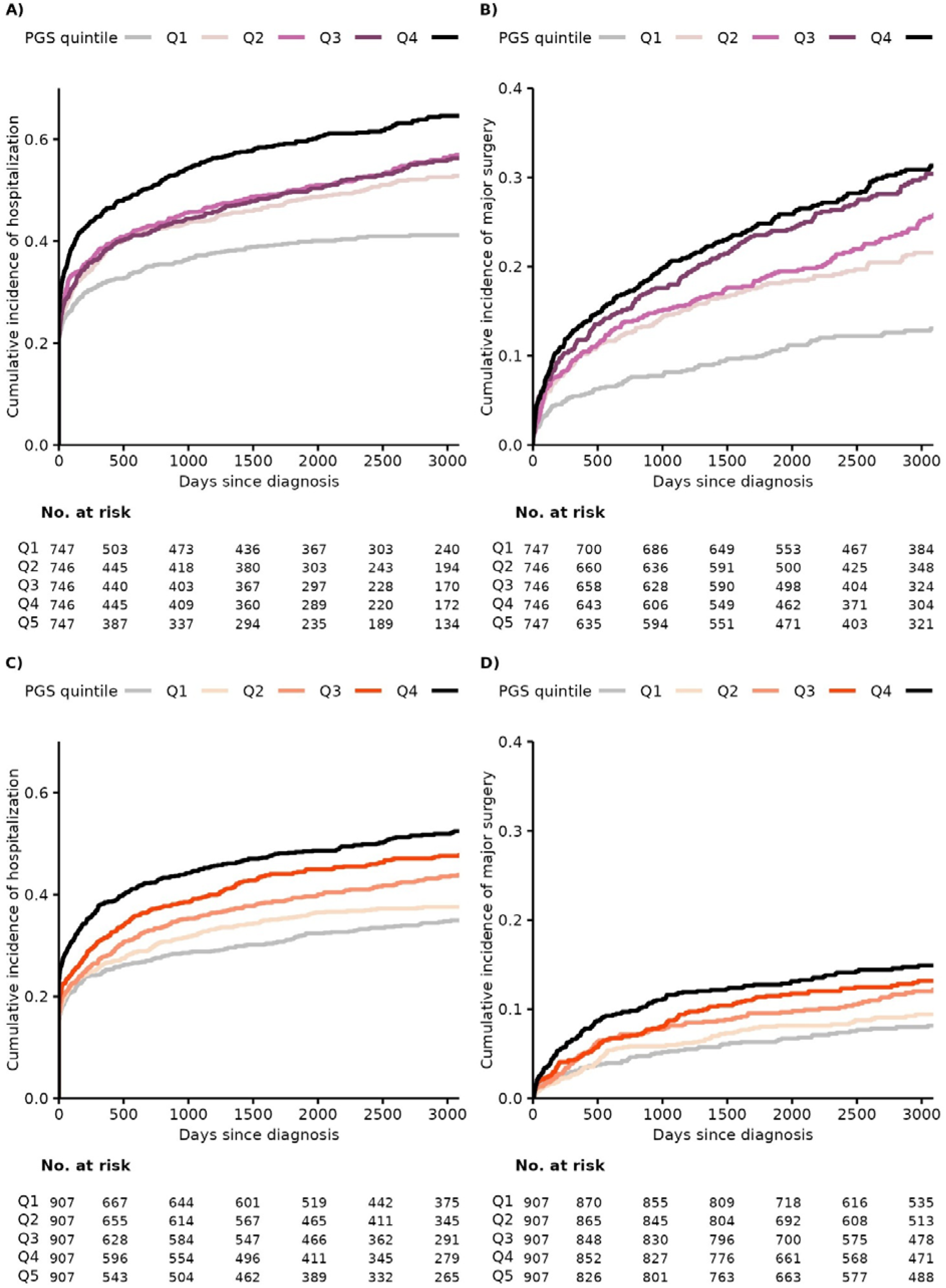
Time to severity event split by PGS quintiles. **A)** Cumulative incidence of IBD-related hospitalization split by CD susceptibility PGS quintiles within the CD cohort. **B)** Cumulative incidence of IBD-related major surgery split by CD susceptibility PGS quintiles within the CD cohort. **C)** Cumulative incidence of IBD-related hospitalization split by UC susceptibility PGS quintiles within the UC cohort. **D)** Cumulative incidence of IBD-related major surgery split by UC susceptibility PGS quintiles within the UC cohort. UC: ulcerative colitis, CD: Crohn’s disease, PGS: polygenic score, Q1-Q5: Quintiles 1 to 5, where Q1 has the lowest quintile of PGS scores, and Q5 the highest.

We generated a binary classification of severe and less severe disease courses based on the first three years after diagnosis, as time to the first event does not necessarily reflect the actual severity. We thus subset the CD and UC cohort to patients with at least three years of follow-up after diagnosis (CD=3688, UC=4491) and generated a composite outcome involving number of IBD-related hospitalizations exceeding two days, IBD-related major surgeries and treatment courses with systemic corticosteroids. 1164 patients with CD (31.6%) were classified as having a severe disease course. Correspondingly, 1224 patients with UC (27.3%) were classified as having a severe disease course.

The PGS of IBD susceptibility was significantly associated with severe versus less severe disease for both CD and UC (Figure 3A-B). Patients with severe CD had higher PGS than the remaining patients with CD (odds ratio [OR]=1.25, 95% CI =1.16-1.35 per SD increase). Accordingly, 39.2% of CD patients in the highest PGS quintile experienced a severe disease course compared to 23.6% of CD patients in the lowest PGS quintile. This was also the case for patients with UC (OR =1.33, 95% CI=1.24-1.43 per SD increase), where 34.9% of patients in the highest PGS quintile experienced a severe disease course compared to 20.8% in the lowest PGS quintile.

**Figure 3:**
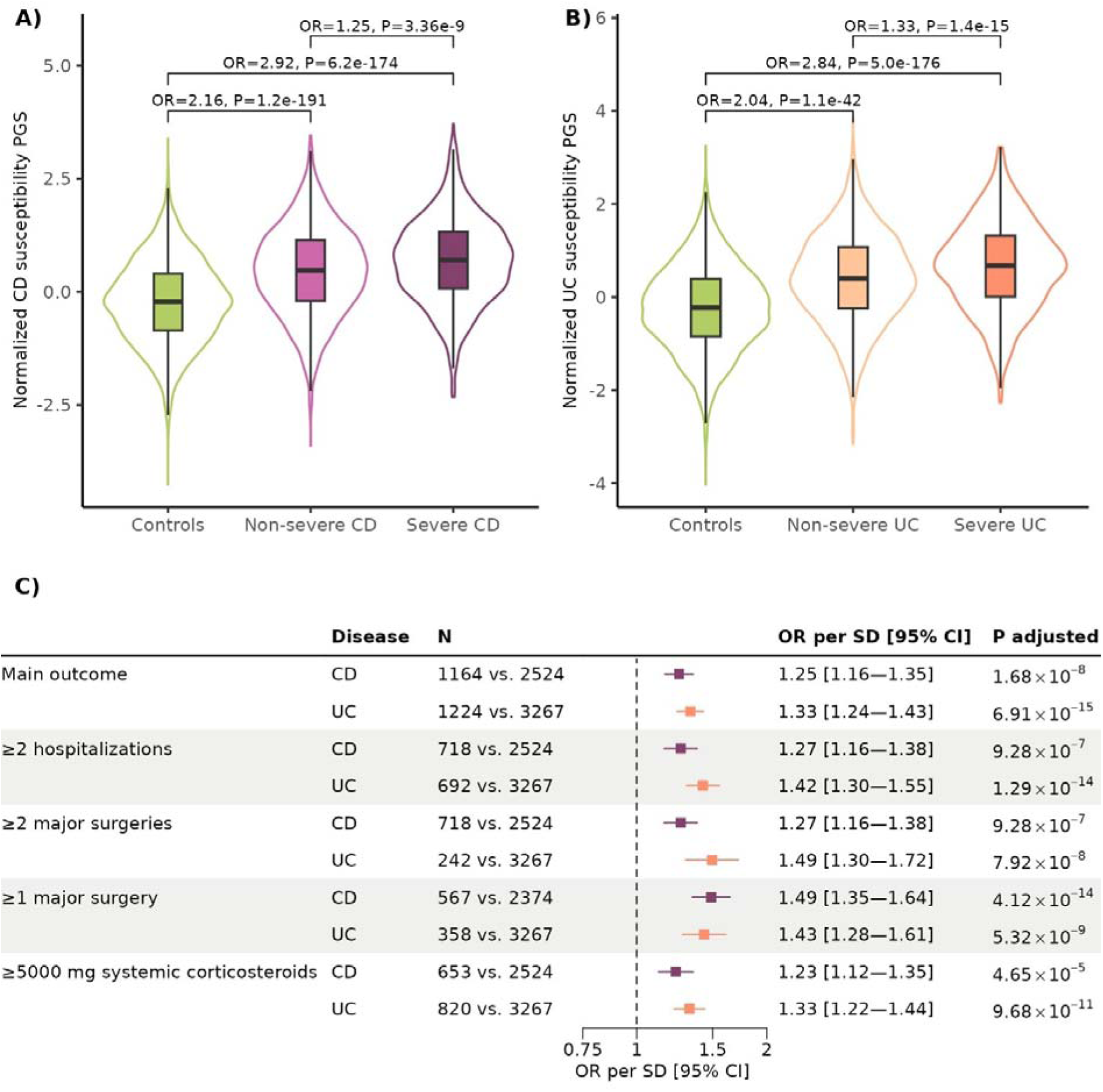
Susceptibility PGS related to IBD severity. **A)** Violin plot of PGS values of CD susceptibility (normalized to SDs from the mean) for controls and CD severity groups. **B)** Violin plot of PGS values of UC susceptibility (normalized to SDs from the mean) for controls and UC severity groups. **C)** Forest plot of the estimated association between the PGS of susceptibility for developing CD and UC and the different entities making up the combined severity definition. All summary statistics are outputted from logistic regressions. *P*-values were adjusted for multiple testing using the Bonferroni correction for ten tests. CD: Crohn’s disease, UC: ulcerative colitis, OR: odds ratio, PGS: polygenic score, SD: standard deviation, CI: confidence interval

We investigated the association between susceptibility PGS and the secondary severity outcomes within the follow-up period: two or more IBD-related hospitalizations exceeding two days, two or more IBD-related major surgeries, one or more IBD-related major surgeries, and a total use of at least 5000 mg systemic corticosteroids (Figure 3C). In general, we observed that all individual components of the main outcome were associated with susceptibility PGS with a consistent effect size ranging from 1.23-1.49 and with overlapping CIs.

### Susceptibility PGS is associated with treatment patterns

We further investigated the association between susceptibility PGS and treatment patterns (Figure 4). For this, we focused the analysis on patients diagnosed since 2003 and with three years of follow-up (CD=3353, UC=4149). For both patients with CD and UC we observed significantly increased use of both biologics (CD: OR=1.35, 95% CI=1.25-1.45, UC: OR=1.31, 95% CI=1.21-1.42 per SD increase) and immunomodulators (CD: OR=1.47, 95% CI=1.36-1.58, UC: OR=1.29, 95% CI=1.20-1.38 per SD increase) within the first three years following diagnosis. We further observed a higher proportion of patients administered several types of biologic therapy and immunomodulator with higher PGS quintile, as well as having multiple treatments with systemic corticosteroids.

**Figure 4:**
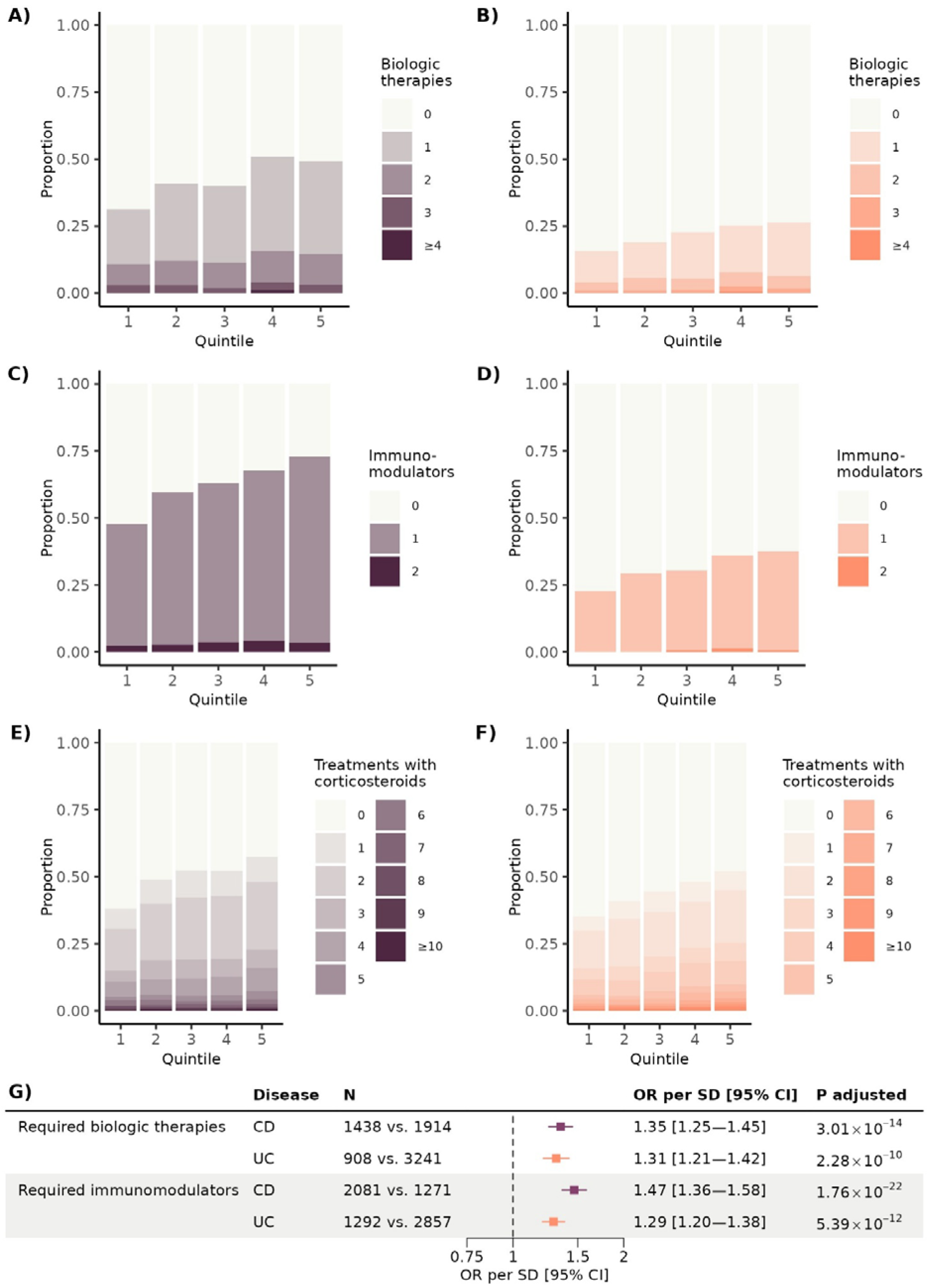
Susceptibility PGS related to treatment use. Per quintile of susceptibility PGS, we generated the number of different biologic therapies used in the first three years after diagnosed based on the proportion of patients with CD (**A**) and UC (**B**). Similarly, we generated the number of different immunomodulators used in the first three years after diagnosed based on the proportion of patients with CD (**C**) and UC (**D**). Lastly, we calculated the total number of treatments with systemic corticosteroids (one treatment is defined as 1500 mg prednisolone equivalent dose) that patients received in the three years after diagnosis based on their PGS quintile for patients with CD (**E**) and UC (**F**). CD: Crohn’s disease, UC: ulcerative colitis, PGS: polygenic score, OR: odds ratio, SD: standard deviation, CI: confidence interval

### Disease extent as a mediator of CD severity

We hypothesized that disease extent could be mediating the observed association between susceptibility PGS and the severity of the disease course in CD and UC. Thus, we conducted sensitivity analyses based on a susceptibility PGS calculated excluding the MST1, NOD2, ATG16L1, and HLA regions, known to associate with disease extent in IBD.^9,21^ The estimated HR comparing time to IBD-related hospitalization exceeding two days for highest versus lowest PGS quintile was thus 1.54 (95% CI=1.33-1.78) for patients with CD, and 1.77 (95% CI=1.54-2.05) for patients with UC (Figure S4). Equivalent results for time to major IBD-related surgery were HR=2.32 (95%CI=1.86-2.91) for patients with CD and HR=1.74 (95% CI=1.32-2.29) for patients with UC. Binary analyses likewise reduced the estimated effect sizes, however the associations remained statistically significant (Figure S5-S6).

Lastly, for the patients with available pathology registrations in the interval from 60 days before diagnosis until three years after (CD=1970, UC=3067), we inferred disease extent. Including disease extent as covariates in the statistical analyses reduced the estimated association, especially for CD (Figure 5). Time to IBD-related hospitalization exceeding two days and to major IBD-related surgery, as well as the OR of experiencing at least one major IBD-related surgery and requiring treatment with biologic therapies remained statistically significant in patients with CD after taking disease extent into account. In patients with UC, all outcomes remained statistically associated with susceptibility PGS after adjusting for disease extent.

**Figure 5:**
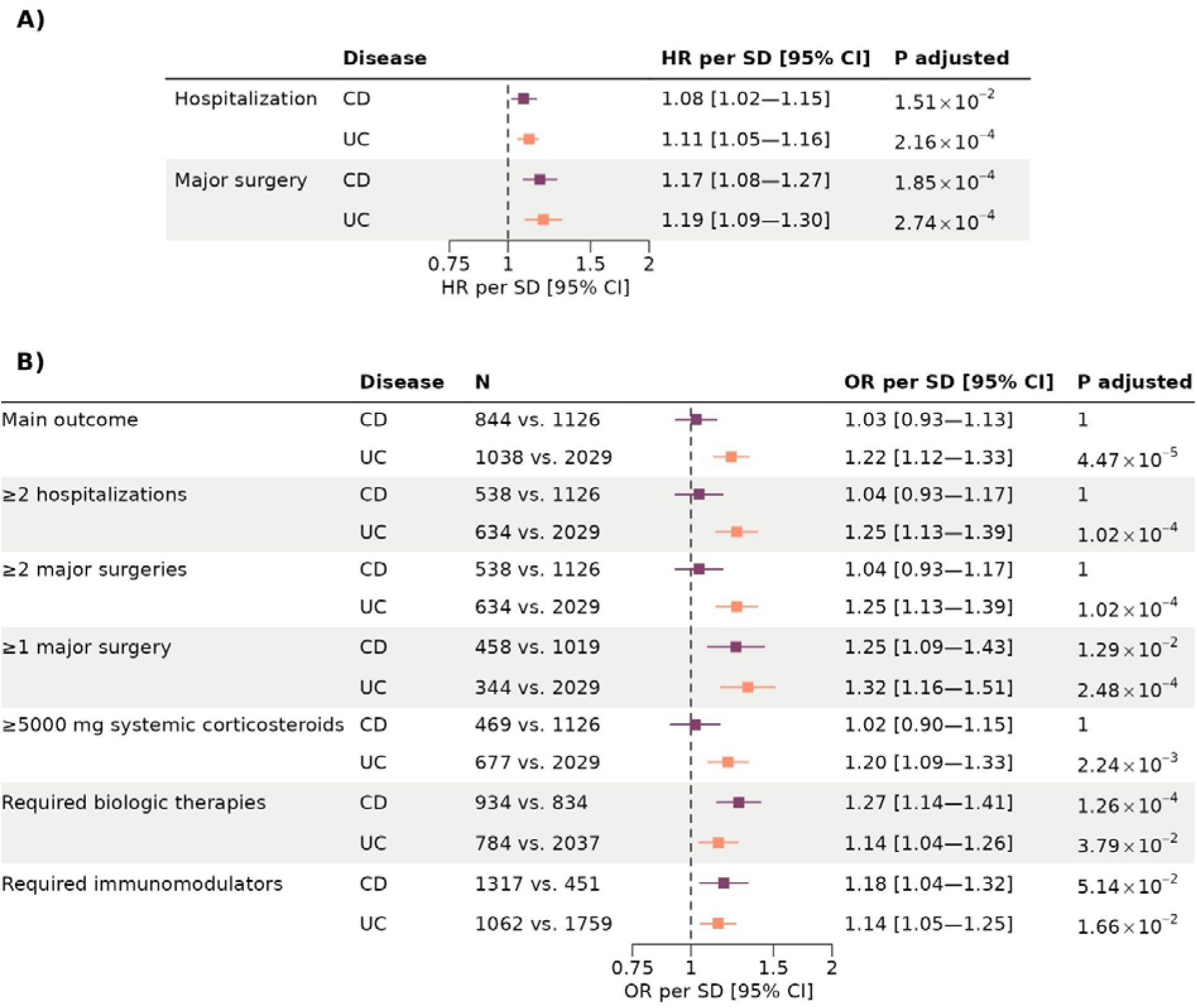
Results of sensitivity analyses adjusting for disease location and extent. We inferred disease location (CD) and extend (UC) from the registries and adjusted for them as sensitivity analyses of the association between susceptibility PGS and disease outcomes. **A**) The estimated HR for time to hospitalization and major surgery, also adjusting for disease location and extend. **B**) All binary disease outcomes were likewise adjusted for disease location and extend. CD: Crohn’s disease, UC: ulcerative colitis, PGS: polygenic score, OR: odds ratio, SD: standard deviation, CI: confidence interval, HR; hazard ratiow

## Discussion

In this population-wide study of more than 8000 patients with IBD with available GWAS data and long-term follow-up, we investigated the influence of IBD susceptibility genetics on the course of IBD using known susceptibility PGS for IBD and a range of longitudinal severity outcomes, including hospitalizations, major surgeries, medication use, and results from biochemistry laboratory tests. We found an association between the aggregated genetic susceptibility to CD and UC and disease severity, however, the association differed between CD and UC, as it appeared to be mainly driven by disease extent in CD, whereas no such pattern was observed in UC.

Using longitudinal nationwide data from biochemistry laboratory tests, we showed that the IBD susceptibility PGS associated with more gastrointestinal inflammation in the form of higher values of f-calpro at time of diagnosis of both CD and UC and during follow-up in patients with CD. Furthermore, susceptibility PGS associated with lower hemoglobin at diagnosis of CD and UC, and elevated CRP in CD but not in UC.

When splitting time to hospitalization exceeding two days and major surgery by PGS quintiles, we observed an additive pattern for both CD and UC, where a higher genetic susceptibility to CD and UC associated with a proportionally higher hazard of hospitalization and major surgery.

In patients with CD, the susceptibility PGS associated most strongly with undergoing major surgery and needing treatment with biologic therapies or immunomodulators. In patients with UC, the susceptibility PGS associated strongly with major surgery and multiple hospitalization events, as well as with a need of treatment with systemic corticosteroids, immunomodulators, and biologic therapies.

Three important susceptibility genes (MST1, NOD2, ATG16L1 and HLA) have been identified to associate with disease extent. Recalculating the PGS without these regions highly reduced the estimated associations with disease course outcomes for patients with CD. Thus, it is possible that the link between susceptibility genetics and disease course severity is mediated by disease extent. It was however not the case for patients with UC, where associations between susceptibility PGS and disease severity were mainly unaltered by adjustment for disease extent. Cleynen *et al.* showed that excluding the three genetic loci from the PGS generation was not sufficient to remove the association with IBD phenotypes. Thus, on the subset of patients with available pathology, we inferred disease extent and included those as covariates in sensitivity analyses. For patients with CD, the adjustment for disease extent highly reduced the estimated association between susceptibility PGS and disease outcomes, which is much in line with the previous findings from Lee et al.^7^ It should, however, be noted that selecting patients with available pathology registrations increased the proportion of patients with severe CD from 31.6% to 42.8%, which may introduce selection bias and explain the diminished association. Also, some outcomes remained statistically significant in CD, including those for major surgeries and exposure to biologic therapies and immunomodulators, after adjusting for extent. Notably, and hitherto unstudied at the population-level, an increased risk of UC associated with an increased risk of experiencing a severe disease course independent of the genetic susceptibility of disease extent. This is much in line with our previous findings, that the susceptibility allele HLA-DRB1*01:03 also greatly associated with severity in UC.^18^

Complementing the findings from Lee *et al.* and Cleynen *et al.,* our results suggest that IBD susceptibility genetics only explain part of the severity of IBD, hence leaving room for other factors at play. These should be investigated together with genetics and included in future prognostic prediction models. This observation is promising, as it points towards the possibility of modulating the effects of genetic susceptibility through lifestyle interventions directed towards potential environmental factors that are yet to be uncovered. Future prediction models of IBD severity may include the gut microbiota profile^23,24^ and biochemistry^25^ in addition to genetics. The present study is hopefully a step towards the development of algorithms based on genetics, environmental exposures, and molecular pathways which in the future can identify patients in need of increased monitoring and intensified treatment to prevent disease from progressing and improving their quality of life.

The primary strength of this study is its nationwide character and unselected nature, which assures generalizability of findings, as well as the complete longitudinal follow-up of patients. Identifying IBD patients using the Danish registries has previously been validated with a positive predictive value of 95%^26^ and completeness of 75-94%.^26,27^ A further strength is the use of unique population-based samples for genetic analyses, which was combined with lifetime health data.

Our study also has potential limitations, which need to be considered. The cohort is rather young, and our results may primarily reflect the course of IBD in this part of the patient population. Also, inferring disease extent from the national registries are at risk of introducing bias in cohort selection, as mentioned previously, and as the prediction of disease extent has an estimated positive predictive value ranging from 0.65 to 0.85 depending on the disease extent category^22^. Laboratory analyses were not available for all patients and their availability may potentially reflect severity. However, to our knowledge no study has been able to test all these parameters over time in a cohort where genetics were also available.

## Conclusion

This study presents novel evidence of an association between the genetic susceptibility burden and subsequent severity of IBD over time, which differs between CD and UC. Patients with higher susceptibility PGS have increased f-calpro and CRP levels, and decreased hemoglobin levels. They experience more hospitalizations, major surgeries and medical treatments, suggesting that genetics of disease susceptibility and disease severity are not orthogonal. However, for patients with CD these associations were greatly mediated by disease extent which was not observed for UC. Our findings of a dose-response relationship between the genetic burden of IBD susceptibility and disease severity provide a new biological understanding of disease severity.

## Acknowledgements

We thank the nurses at the outpatient clinic at the Department of Gastroenterology and Hepatology at Aalborg University Hospital, Denmark, for their important contribution to the sample collection. We thank Prof. James C Lee for helpful discussions on result interpretations and validity.

## Conflict of Interest

TJ reports consultancy for Ferring and Pfizer. LL reports speaker and consultancy fee from Takeda and Eli Lilly advisory board for Tillotts, Eli Lilly, Abbvie, and Celltrion. HAJ reports speaker fee for Tillotts. The remaining authors have no disclosures.

## Author contributions

Conceptualization: MVV, KHA, AKN, MB, AS, TJ

Methodology: MVV, KHA, AKN, HSC, RFB, MB, AS, TJ

Software: MVV, JVTL, AKN, AS

Formal Analysis: MVV, AKN, JVTL, HSG, AS

Investigation (GWAS analysis): MBH, JBG

Resources: LL, HK, KRN, TJ

Data Curation: JVTL, AKN, MVV

Writing – Original Draft: MVV, KHA

Writing – Reviewing & Editing: All authors

Visualization: MVV

Supervision: AS, KHA, TJ

Project Administration: TJ

Funding Acquisition: LL, AS, TJ

## Funding

Danish National Research Foundation, DNRF148, Lundbeck Foundation, R313-2019-857, Novo Nordisk Foundation, NNF23OC0087616, The Colitis-Crohn Foreningen.

## Data and code availability

This work uses data from the Danish National Health registries (https://sundhedsdatastyrelsen.dk), which is protected by the Danish Act on Processing of Personal Data. To access the data an application is required, which must be approved from the Danish Data Protection Agency and the Danish Health Data Authority.

Code will be made available: https://github.com/marievibeke/PGS_severity

## Notes

### Summary of Updates

The manuscript has been revised by including a sensitivity analysis. This sensitivity analysis evaluates disease location as a possible mediatior of association.

