## Supplementary material for "Genetic risk of inflammatory bowel disease is associated with disease course severity"

**Supplementary online content:**

**Supplement 1: Genotyping and quality control information**

**References**

**Table S1**

**Table S2**

**Figure S1**

**Figure S2**

**Figure S3**

**Figure S4**

**Figure S5**

**Figure S6**

**Supplement 1: Genotyping and quality control**

DNA was extracted from the neonatal blood spots in the PREDICT neonatal blood spot cohort (PREDICT-NBS cohort) and from full blood samples in the Northern Denmark IBD cohort (NorDIBD cohort). Both types of blood samples were extracted using standard protocols. Extracted DNA was genotyped using a modified version of the global screening array v.3 from Illumina (Illumina, San Diego, CA, USA), excluding variants with minor allele frequency (MAF) <0.01 and excluding variants within actionable genes (based on the American College of Medical Genetics). Genotype calling was carried out by using Gentrain V3 from GenomeStudio. A total of 21,233 unique samples were genotyped.

Quality control (QC) was conducted using Plink[1,2] and BCFtools.[3] All single nucleotide positions (SNPs) were converted to the hg19 forward strand and consistency was evaluated by merging with the 1000 Genomes Phase 3 reference panel (1000G).[4] Exclusion steps at genetic variant level were as follows: MAF <0.01, missingness >0.05, Hardy–Weinberg equilibrium p-value <10^-5^, SNPs with unknown position, tri-allelic SNPs, duplicated SNPs, and SNPs not matching the 1000G reference panel. Exclusion steps at individual level were: Individuals with missingness >0.05, heterozygosity rate outside the mean ± three standard deviations (SD), and where their phenotypic sex did not match the genotypic sex estimated by Plink.

A total of 19,680 individuals and 470,194 SNPs passed QC.

Phasing was conducted using Shapeit4 v4.1.3.[5] Imputation was conducted using Minimac4 v1.0.2 to the 1000G v3.0.0.[6] The imputed dataset consisted of 48,935,166 variants. Post-imputation QC included removal of SNPs with MAF <0.01 and imputation score (R2) <0.3.

The final set of SNPs used for the analysis included a total of 9,518,572 variants.

We imputed HLA alleles using the HIBAG package[7] at the two-field (4-digit) resolution paired with the multi-ethnic model for the Illumina Infinium Global Screening Array v2.0 supplied by the package authors.

Later evaluations of the cohorts revealed monozygotic twins based on genetics, but with different parents and/or dates of birth based on information from the Danish registers. These specific register-conflictive individuals were removed. Lastly, we estimated identity-by-decent for all pairs of individuals using Plink and excluded related individuals (pi-hat ≥0.1875) across the two cohorts from the largest cohort (the PREDICT-NBS cohort).

We only included individuals with European ancestry in the analyses. We projected our samples into the space of the pre-calculated principal component coordinates for the 1000G samples using the variant loadings provided by the AKT toolkit’s authors.[8] We then trained a random forest model on the 1000G sub-cohort labels (European, admixed American, South Asian, East Asian, African) and used the classifier to predict the membership for our samples (minimum probability = 0.9). Most of our samples (18,665; 94.8%) were classified as Europeans.

We reapplied a filter of MAF ≥0.01 after selecting the study populations.

We calculated principal components (PCs) to include as covariates to adjust for population structure in the models. The PCs were calculated using a set of common variants excluding long-range linkage disequilibrium regions. The PCs were generated based on unrelated individuals and variant loadings were then projected onto the full dataset.

**Table S1: Definition and relevant codes for included variables.**

| **Variable** | **Definition and codes** | | **Source** |
| --- | --- | --- | --- |
| **IBD-related hospitalization** | Inpatient contact with A-diagnosis of  ICD-10: K50/K51 or B-diagnosis of K50/K51 and A-diagnosis of one the following | | Danish National  Patient Registry |
| Abdominal pain | ICD-10: R100, R101, R102C, R103 | | Danish National  Patient Registry |
| Nausea and vomiting | ICD-10: R11 | |  |
| Non-infectious gastroenteritis | ICD-10: K529 (excl. K529B1) | |  |
| Rectal- or anal bleeding | ICD-10: K625 | |  |
| Fistula | ICD-10: K603-605, K316E, K632,  N822-825, N828F | |  |
| Abscess | ICD-10: K610-14, K630, K650A,  K650G, K650H, L023C | |  |
| Stenosis | ICD-10: K264, K566+F+G | |  |
| Ileus and sub-ileus | ICD-10: K566C, K567 | |  |
| **IBD-related major surgery** |  | |  |
| Intestinal resections | NCSP: KJGB, KJFB (excl. KJFB10+13) | | Danish National  Patient Registry |
| Enteroenterostomy | NCSP: KJFC | |  |
| Enterostomy | NCSP: KJFF | |  |
| Colectomy | NCSP: KJFH | |  |
| Intestinal stricture-plasty | NCSP: KJFA60, KJFA61, KJFA63 | |  |
| Other local intestinal surgery | NCSP: KJFA96, KJFA97 | |  |
| Other intestinal surgery | NCSP: KJFW | |  |
| Stenosis surgery without  resection or adhesiolysis | NCSP: KJFL | |  |
| **Systemic corticosteroids** | ATC: H02AB01, H02AB02, H02AB04, H02AB06, H02AB07, H02AB08, H02AB09 | | Danish National Prescription Registry |
| **Biologic therapies** |  | |  |
| TNF-α inhibitor | C_OPR: BOHJ18A | | Danish National Prescription Registry |
| Infliximab | C_OPR: BOHJ18A1 | |  |
| Adalimumab | C_OPR: BOHJ18A3 | |  |
| Golimumab | C_OPR: BOHJ18A4 | |  |
| Certolizumab | C_OPR: BOHJ18A5 | |  |
| Ustekinumab | C_OPR: BOHJ18B3 | |  |
| Vedolizumab | C_OPR: BOHJ19H4 | |  |
| Natalizumab | C_OPR: BOHJ26 | |  |
| Risankizumab | C_OPR: BOHJ19N1 | |  |
| **Immunomodulators** |  | |  |
| Azathioprine | ATC: L04AX01; C_OPR: BWHB83 | | Danish National Prescription Registry |
| Mercaptopurine | ATC: L01BB02 | |  |
| Methotrexate | ATC: L04AX03/L01BA01; C_OPR: BWHA115 | |  |
| **Biochemistry tests** |  | |  |
| CRP | NPU19748, NPU01422 | | The Danish nationwide Register of Laboratory Results for Research |
| Fecal calprotectin | NPU19717, NPU26814 | |  |
| Hemoglobin | NPU02319 | |  |
| **Disease location/extend** |  | |  |
| L1 (ileal) | T64000, T64020, T64040, T64120, T65200, T65210, T65300, T65310, T65905 | M42100, M43000, M43005, M43009, M43030, M44001, M44002, M44200, M44202 & S6214, S6216 | Danish Pathology Registry |
| L2 (colonic) | T67005, T67010, T67011, T67012, T67100, T67105, T6710C, T67200, T67205, T67210, T67300, T67305, T67310, T67400, T67405, T67410, T67500, T67505, T67510, T67600, T67605, T67610, T67700, T67705, T67710, T67920, T67921, T67925, T67965, T67966, T67995, T67996, T68000, T68005, T68010 |  |  |
| L3 (ileocolonic) | T65900, T65925, T65926, T65901, T65902, T67924 |  |  |
| L4 (upper) | T62000, T62010, T62050, T62910, T62911, T62915, T63000, T63010, T63140, T63300, T63310, T63400, T63410, T63500, T63510, T63520, T63530, T63600, T63610, T63620, T63630, T63700, T63910, T63911, T63920, T63930, T63950, T63951, T64300, T64310, T64311, T64312, T51000, T51020, T51030, T51140, T51300, T52110, T52210, T52250 |  |  |
| E1 (proctitis) | T68000, T69000, T69110 | M42100, M43000, M43005, M43009, M43030 & S62140, S62550, S62820 |  |
| E2 (left-sided) | T67600, T67700, T67921, T67995 |  |  |
| E3 (extensive) | T64000, T65100, T65200, T65300, T65900, T67000, T67100, T67200, T67300, T67400, T67500, T67965, T67966 |  |  |

UC: Ulcerative colitis, ICD: International Classification of Diseases, NCSP: Classification of Surgical Procedures, ATC: Anatomical Therapeutic Chemical, C_OPR: operation code, NPU: Nomenclature for Properties and Units, CRP: C-reactive protein

**Table S2: Software, packages, versions, and settings.** Reported settings are those deviating from default.

| **Description** | **Function** | **Package (for R)** | **Version** | **Settings** |
| --- | --- | --- | --- | --- |
| PCA: LD pruning | plink2 |  | 2.00a3.7LM | --indep-pairwise 500 5 0.2 |
| PCA | plink2 |  | 2.00a3.7LM | --freq --pca allele-wts |
| Calculate PGS | plink2 |  | 2.00a3.7LM | --score  cols=scoresums |
| Logistic regression | glm | stats | 4.3.0 | family=”binomial” |
| Cox proportional hazards regression model | coxph(Surv()) | survival | 3.5.8 |  |
| Plotting | ggplot | ggplot2 | 3.5.0 | geom_violin  geom_boxplot  geom_col  geom_point  geom_errorbar |
| Forest plots | forest | forestploter | 1.1.2 |  |
| Survival plots | ggsurvplot | survminer | 0.4.9 |  |
| Combining plots | ggarrange | ggpubr | 0.4.0 |  |

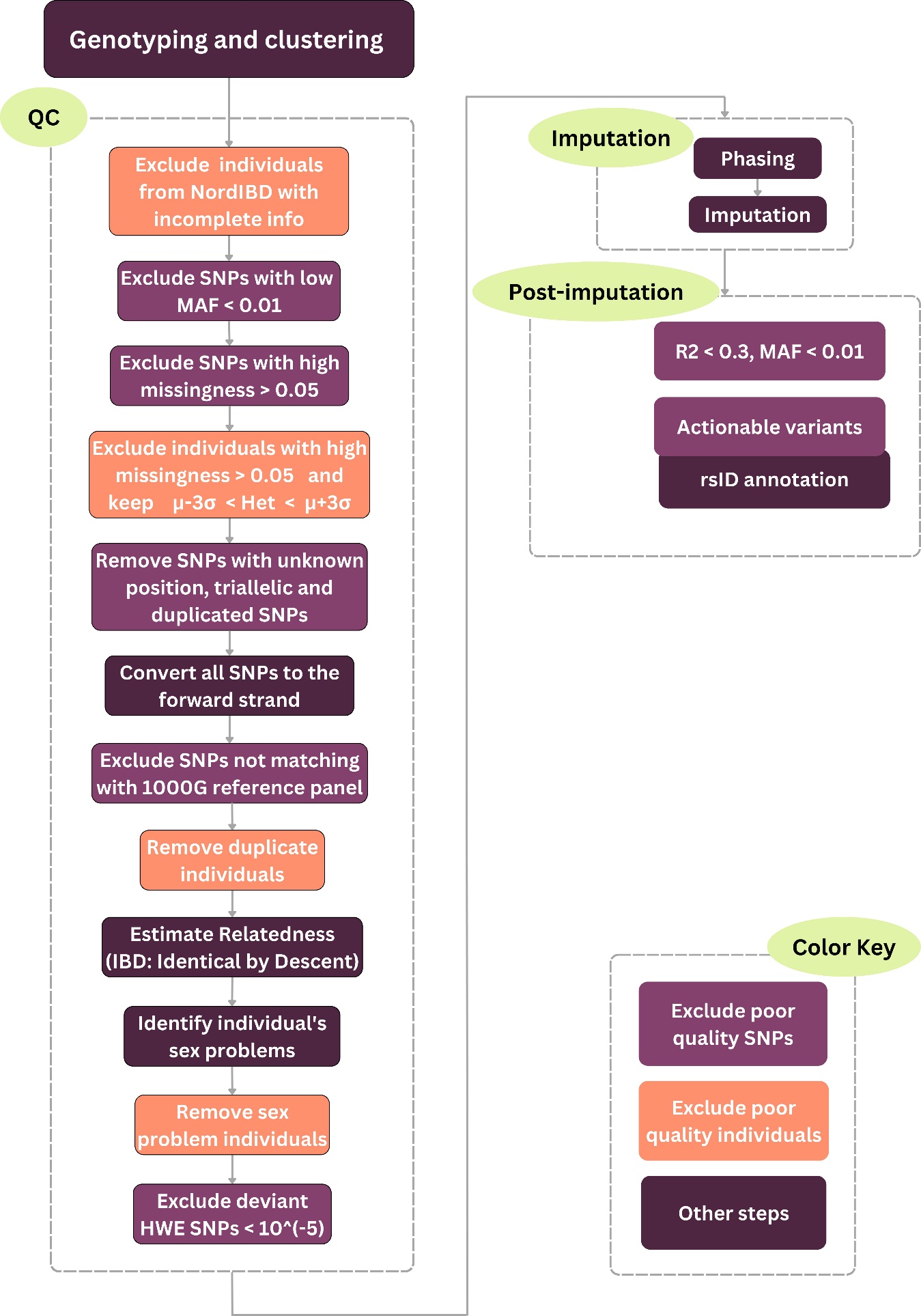

**Figure S1:** Flowchart of the steps included in the genetic quality control, phasing, and imputation of data.

QC: quality control, SNP: single nucleotide position, MAF: minor allele frequency, HWE: Hardy-Weinberg equilibrium

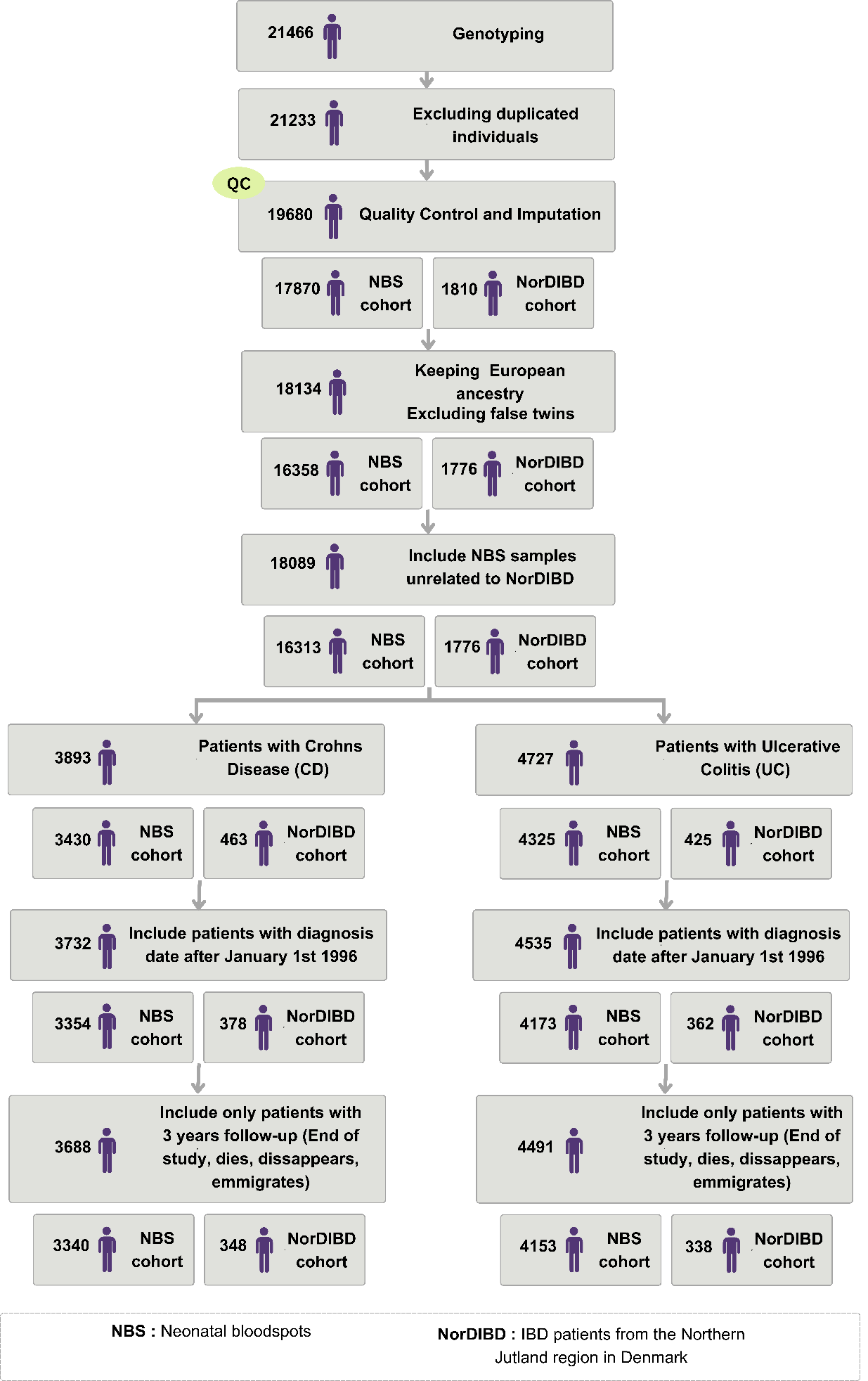

**Figure S2:** A. Flowchart of selection for inclusion of study participants.

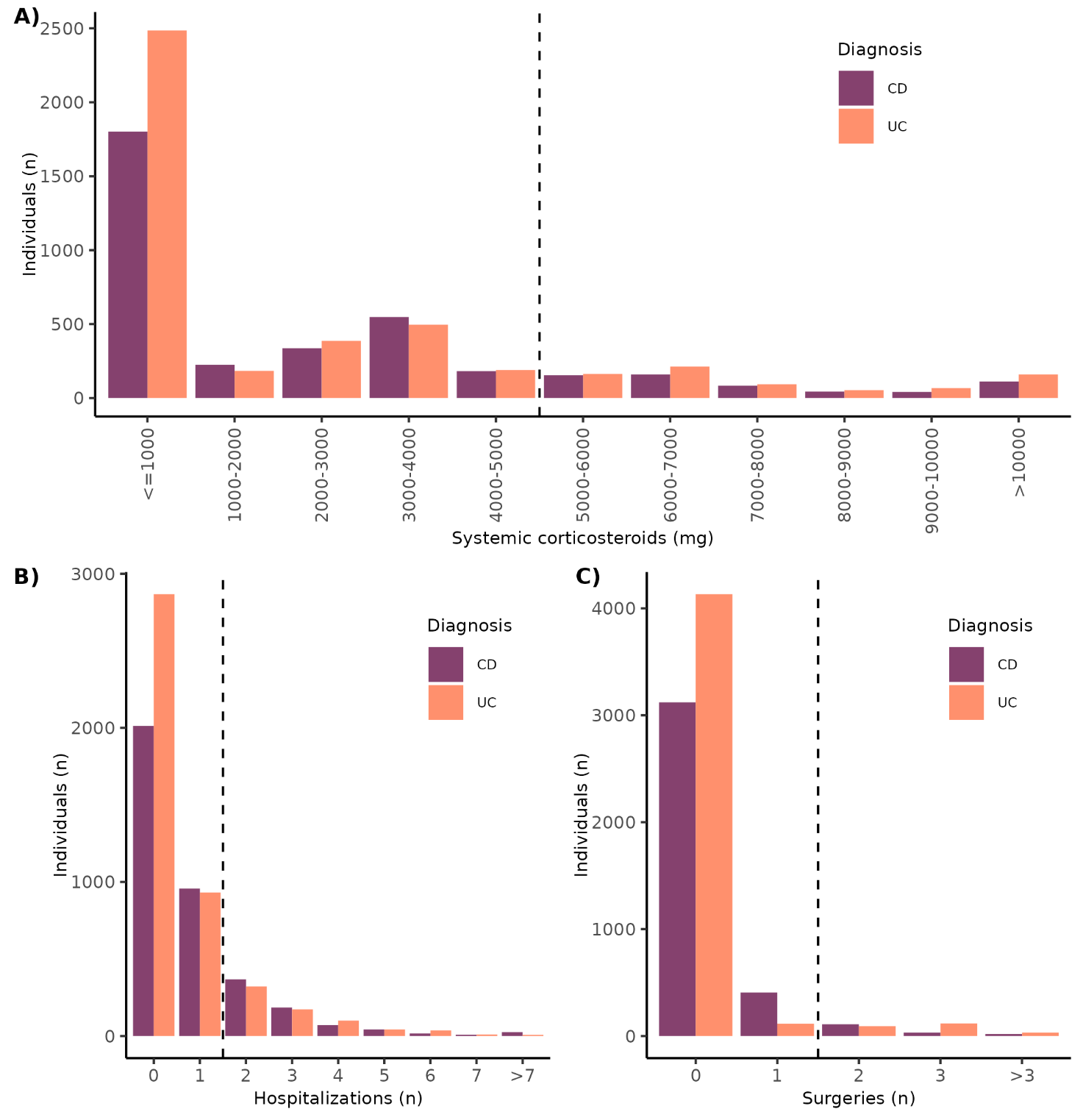

**Figure S3:** **Number of individual outcomes defining disease severity.** Bar plots showing **A)** total use of systemic corticosteroid (mg), **B)** total number of IBD-related hospitalizations exceeding 2 days, and **C)** total number of IBD-related major surgeries in the three years following diagnosis for patients with CD and UC separately. The black dashed line shows the cut-off for defining a severe disease course for CD. For UC, the cutoff for major surgeries was moved to ≥1.

CD: Crohn’s disease, UC: ulcerative colitis

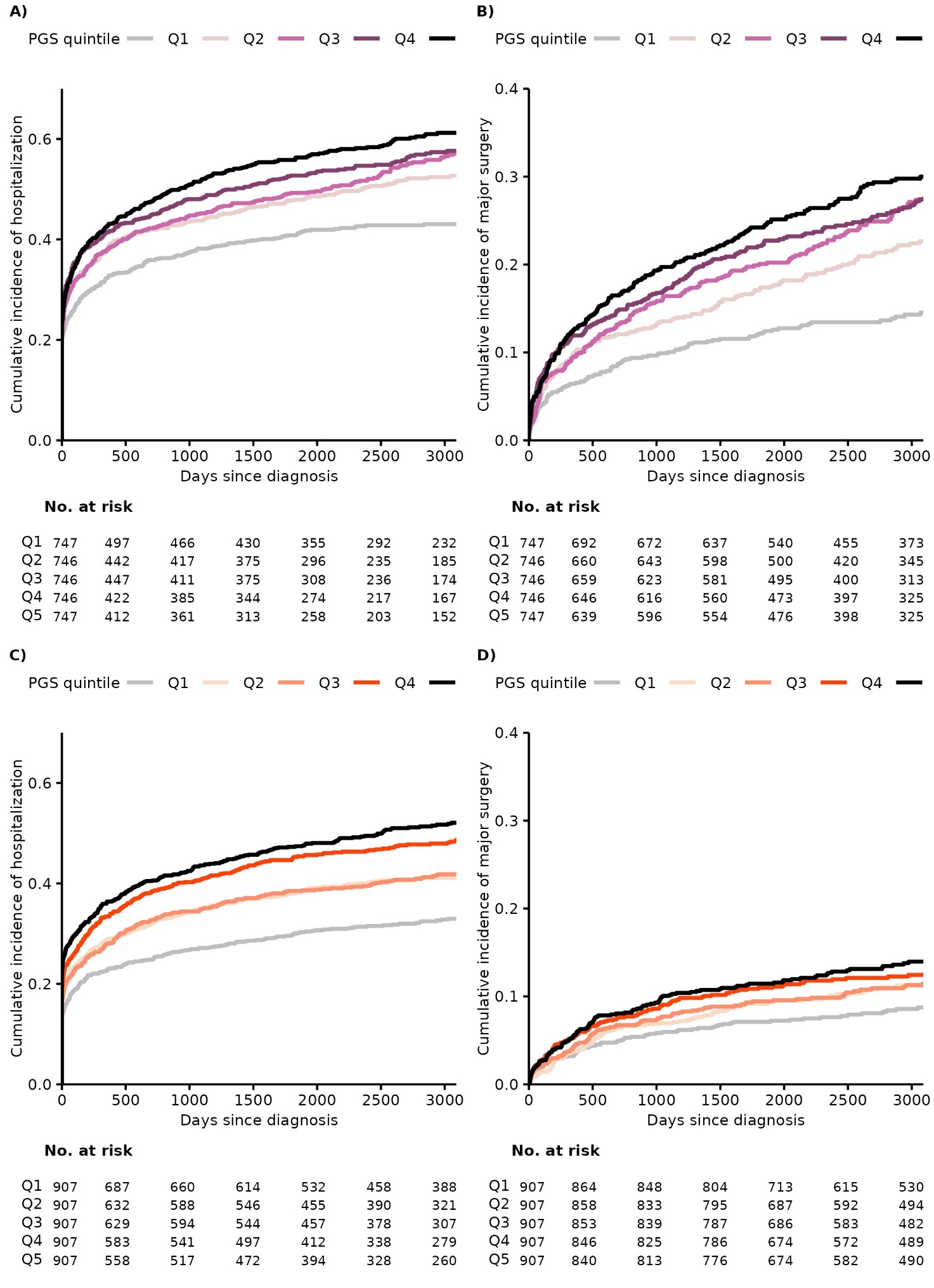

**Figure S4: Time to severity event split by PGS quintiles calculated without genetic loci associated with disease location. A)** Cumulative incidence of IBD-related hospitalization split by CD susceptibility PGS quintiles within the CD cohort. **B)** Cumulative incidence of IBD-related major surgery split by CD susceptibility PGS quintiles within the CD cohort. **C)** Cumulative incidence of IBD-related hospitalization split by UC susceptibility PGS quintiles within the UC cohort. **D)** Cumulative incidence of IBD-related major surgery split by UC susceptibility PGS quintiles within the UC cohort.

UC: ulcerative colitis, CD: Crohn’s disease, PGS: polygenic score, Q1-Q5: Quintiles 1 to 5, where Q1 has the lowest quintile of PGS scores, and Q5 the highest.

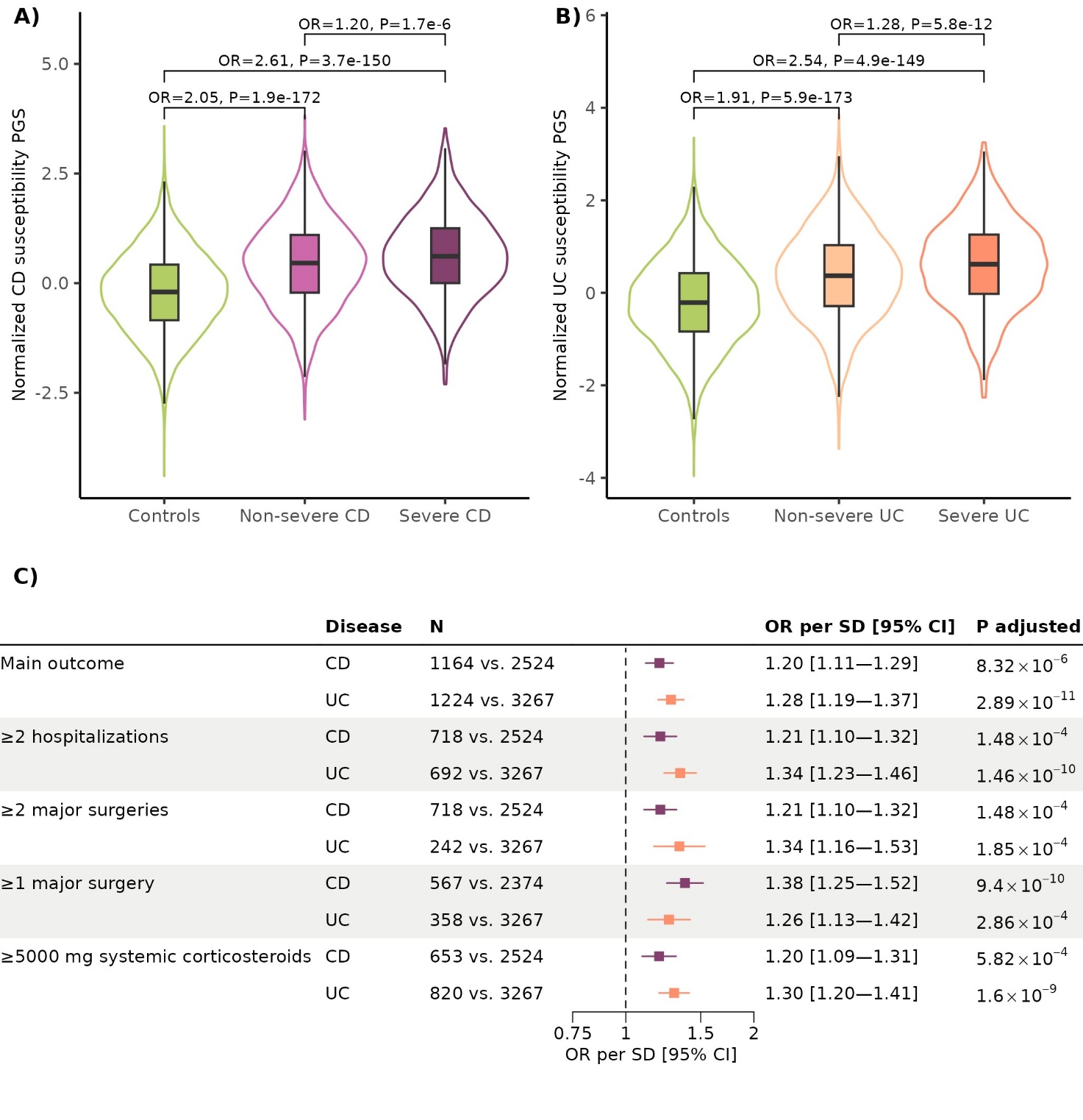

**Figure S5: Susceptibility PGS related to IBD severity, where the PGS was** **calculated without genetic loci associated with disease location. A)** Violin plot of PGS values of CD susceptibility (normalized to SDs from the mean) for controls and CD severity groups. **B)** Violin plot of PGS values of UC susceptibility (normalized to SDs from the mean) for controls and UC severity groups. **C)** Forest plot of the estimated association between the PGS of susceptibility for developing CD and UC and the different entities making up the combined severity definition. All summary statistics are outputted from logistic regressions. P-values were adjusted for multiple testing using the Bonferroni correction for ten tests.

CD: Crohn’s disease, UC: ulcerative colitis, OR: odds ratio, PGS: polygenic score, SD: standard deviation, CI: confidence interval

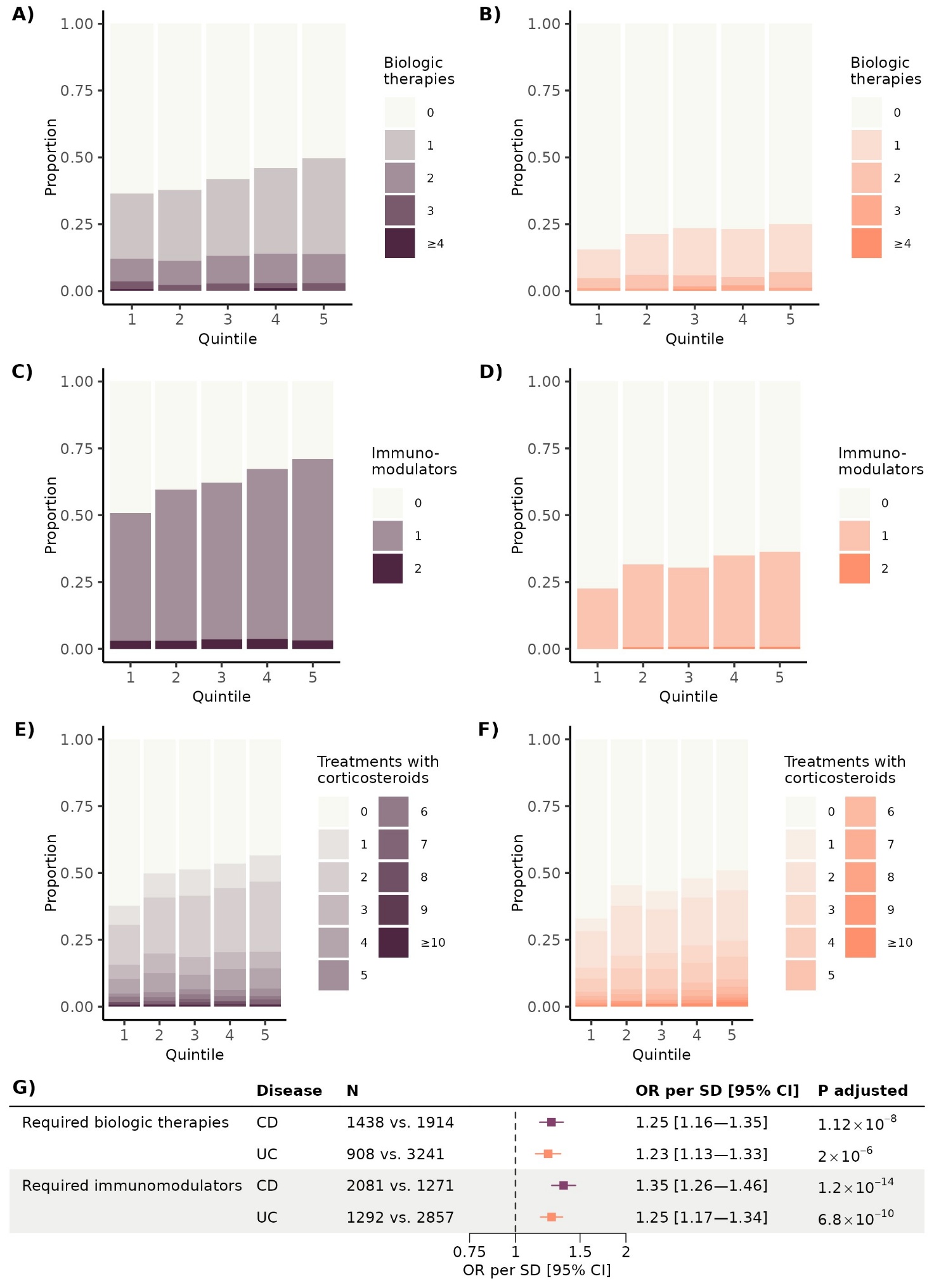

**Figure S6: Susceptibility PGS related to treatment use**, **where the PGS was** **calculated without genetic loci associated with disease location.** Per quintile of susceptibility PGS, we generated the number of different biologic therapies used in the first three years after diagnosed based on the proportion of patients with CD (**A**) and UC (**B**). Similarly, we generated the number of different immunomodulators used in the first three years after diagnosed based on the proportion of patients with CD (**C**) and UC (**D**). Lastly, we calculated the total number of treatments with systemic corticosteroids (one treatment is defined as 1500 mg prednisolone equivalent dose) that patients received in the three years after diagnosis based on their PGS quintile for patients with CD (**E**) and UC (**F**).

CD: Crohn’s disease, UC: ulcerative colitis, PGS: polygenic score, OR: odds ratio, SD: standard deviation, CI: confidence interval
